# Cost-effectiveness of remdesivir and dexamethasone for COVID-19 treatment in South Africa

**DOI:** 10.1101/2020.09.24.20200196

**Authors:** Youngji Jo, Lise Jamieson, Ijeoma Edoka, Lawrence Long, Sheetal Silal, Juliet R.C. Pulliam, Harry Moultrie, Ian Sanne, Gesine Meyer-Rath, Brooke E Nichols

## Abstract

**Background:** South Africa recently experienced a first peak in COVID-19 cases and mortality. Dexamethasone and remdesivir both have the potential to reduce COVID-related mortality, but their cost-effectiveness in a resource-limited setting with scant intensive care resources is unknown.

**Methods:** We projected intensive care unit (ICU) needs and capacity from August 2020 to January 2021 using the South African National COVID-19 Epi Model. We assessed cost-effectiveness of 1) administration of dexamethasone to ventilated patients and remdesivir to non-ventilated patients, 2) dexamethasone alone to both non-ventilated and ventilated patients, 3) remdesivir to non-ventilated patients only, and 4) dexamethasone to ventilated patients only; all relative to a scenario of standard care. We estimated costs from the healthcare system perspective in 2020 USD, deaths averted, and the incremental cost effectiveness ratios of each scenario.

**Results:** Remdesivir for non-ventilated patients and dexamethasone for ventilated patients was estimated to result in 1,111 deaths averted (assuming a 0-30% efficacy of remdesivir) compared to standard care, and save $11.5 million. The result was driven by the efficacy of the drugs, and the reduction of ICU-time required for patients treated with remdesivir. The scenario of dexamethasone alone to ventilated and non-ventilated patients requires additional $159,000 and averts 1,146 deaths, resulting in $139 per death averted, relative to standard care.

**Conclusions:** The use of dexamethasone for ventilated and remdesivir for non-ventilated patients is likely to be cost-saving compared to standard care. Given the economic and health benefits of both drugs, efforts to ensure access to these medications is paramount.

**40-word summary of article’s main point:** The use of remdesivir and dexamethasone for treatment of severe COVID-19 in South Africa is likely to be cost-saving relative to standard care. Enabling access to these medications should be prioritize to improve patient outcomes and reduce total costs.

## Introduction

As of September 2020, there were over 29 million confirmed SARS-CoV-2 infections worldwide, and over 939,000 reported COVID-19 associated deaths.(1) South Africa has the 8th highest number of confirmed cases worldwide with more than 653,000 confirmed infections and 16,000 confirmed COVID-19 deaths.(2) During the surge in cases, intensive care unit (ICU) capacity was reportedly breached in some locations and much-needed oxygen supplies were running low.(3) While increased mortality is associated with several individual-level factors including older age, male sex, and pre-existing comorbidities, systems-level factors such as being admitted to a hospital with fewer ICU beds also play a role.(4) Considering inter-hospital variation in treatment and outcomes, the mortality rate among those in ICU has ranged between 40%-50%.(4-6) To mitigate the mortality impact of COVID-19 in South Africa and other low- and middle-income countries with scant intensive care capacity, efforts to reduce mortality amongst the more severe cases in ICU are paramount. Recently, two therapeutic agents, remdesivir and dexamethasone, have been demonstrated in randomized clinical trials to reduce COVID-related mortality.(7, 8)

Dexamethasone has been shown to decrease mortality by 35% (95% confidence interval [CI]: 18% to 49%) and 20% (95% CI: 8% to 30%) in ventilated ICU patients and non-ventilated patients with oxygen respectively and has shown no clear evidence regarding its impact on recovery time,(8) while remdesivir has been shown to decrease mortality by 30% (95% CI: -4% to 53%) and reduce recovery time from 15 (95% CI: 13-19) to 11 (95% CI:9-12) days amongst ICU patients who are not ventilated.(7) In low- and middle-income countries, not only is ICU capacity limited, but so are budgets for COVID-related care. There has been an additional budget allocation of $1.14 billion for the health response to COVID-19 in South Africa(9), though this budget does not yet include remdesivir or dexamethasone. We therefore conducted a cost-effectiveness analysis to determine the incremental cost per death averted of providing dexamethasone and remdesivir relative to the standard care of critically ill patients in South African ICUs from a health systems perspective. The cost-effectiveness of these regimens is going to become increasingly important for possible eventuality of a COVID-19 resurgence in South Africa, in order to ensure improved ICU capacity and patient outcomes during a future wave.

## Methods

Using the South African National COVID-19 Epidemiology Model (NCEM)(10) and government data on the number of currently available ICU beds in the public and private sector by province(11), we projected the number of people requiring ICU admission and total ICU person days by province by month between August 2020 and January 2021. We then projected costs and COVID-related mortality for four scenarios: 1) dexamethasone administered to ventilated patients (i.e. in an advanced stage of disease illness) and remdesivir to non-ventilated patients (i.e. to those receiving supplemental oxygen as in an earlier stage of disease illness), 2) dexamethasone alone to both non-ventilated and ventilated patients, 3) remdesivir to non-ventilated patients only, and 4) dexamethasone to ventilated patients only; all relative to standard of ICU care (without remdesivir and dexamethasone).

Our effectiveness measure was expressed as COVID-19 deaths averted. The mortality impact for all patients in ICU between August 2020 and January 2021 was estimated by applying the treatment impact of remdesivir and dexamethasone on mortality from the results of recent randomized controlled trials (7, 8) and impact on mortality that remdesivir could have through decreasing the average length of stay of patients (from 15 to 11 days), when ICU capacity is expected to be breached.(7) (Table 1) Based on South African COVID-19 hospitalisation data and treatment guidelines(12), we assumed that 42% of patients in ICU require mechanical ventilation, while the remaining (58%) patients do not yet require mechanical ventilation (though they might require supplementary oxygen). A course of dexamethasone (oral/intravenous administration) is assumed to be 10 days long, and a course of remdesivir (intravenous) is assumed to be five days long.

**Table 1.** Comparison between remdesivir and dexamethasone as COVID-19 treatment in South Africa.

|  |  | Remdesivir | Dexamethasone |
| --- | --- | --- | --- |
| Regimen (14) |  | Antiviral: 200 mg IV loading dose following by 100 mg IV daily for up to 5 days for patients who are hospitalized. | Steroid: 6 mg PO / IV for up to 10 days for patients with an oxygen requirement and/or requiring mechanical ventilation. |
| Clinical trial (7,8) |  | National Institute of Allergy and Infectious Diseases (NIAID) Clinical Trial | Oxford RECOVERY trial |
| Clinical outcome (7,8) | Median time to recovery | From 15 (13–19) days in standard care to 11 (9–12) days with Remdesivir | N/A |
|  | Overall (Hazard ratio) | RR 0.70 (95% CI 0.47–1.04)* | RR 0.83 (95% CI 0.74–0.92) |
|  | Patients not receiving oxygen | RR 1.38 (95% CI 0.94–2.03) | RR 1.22 [95% CI 0.93 to 1.61] |
|  | Patients receiving oxygen | RR 1.47 (95% CI 1.17–1.84) | RR 0.80 [95% CI 0.70 to 0.92]* |
|  | Patients receiving high-flow oxygen or noninvasive mechanical ventilation | RR 1.20 (95% CI 0.79–1.81) |  |
|  | Patients receiving mechanical ventilation | RR 0.95 (95% CI 0.64–1.42) | RR 0.65 [95% CI 0.51 to 0.82]* |
| Availability (31)(32) |  | Possibly in short supply | Currently in market |
| Any adverse events (24, 33) |  | Remdesivir vs placebo: 66% vs. 64% for any group, 8% vs. 14% for patients receiving oxygen | N/A |

\*Asterisk represents the mortality reduction rates used for remdesivir and dexamethasone in the model.

We estimated resource use from the perspective of the provider, the South African healthcare system, for all ICU-days accrued by COVID-19 patients from August 2020 and January 2021 for each scenario. (Note that since COVID-19 patients will be accommodated in both the private and public sector, depending on need and availability, and contracting arrangements towards that end were in place in all provinces at the time of writing, we combined both sectors for the purpose of this analysis.) We calculated health system operational cost (excluding drug costs) based on the latest NCEM outputs (i.e. required total ICU beds and days) with and without remdesivir and/or dexamethasone. We used an ICU cost per person day of $1,128 ($665-$1,172) based on an ingredients-based costing analysis including capital equipment, current South African ICU staffing norms and salaries, and overhead costs representative of South Africa public hospital settings.(13) Capital assets (e.g. ventilators) were annualized based on the relevant years of useful life (e.g. 10 years) and discounted at a 5% annual rate based on the South Africa pharmacoeconomic guidelines.(14)

For the scenarios including novel therapeutics, we assumed the additional cost of a full course of remdesivir ($55 per 100mg IV, corresponding to $330 per patient course)(15) and/or dexamethasone ($3.13 per 4mg, corresponding to $31 per patient course)(16), for the respective scenarios that include these therapeutics. (Table 2) Costs were converted from South African Rands (ZAR) to US dollars (USD) at the average exchange rate over the period August 2020 and January 2021 (1 USD = 16.95 ZAR).(17) Incremental cost effectiveness ratios were calculated as the incremental cost in 2020 USD of each therapeutic scenario over standard care per death averted.

**Table 2.** Key input parameters.

| Patient disease status in ICU | Base | Low | High | Distribution | Source |
| --- | --- | --- | --- | --- | --- |
| % of patients requiring mechanical ventilation | 42% | 34% | 49% | Triangular | (7, 10,11) |
| Days of hospitalization for non-ventilated patients | 15 | 7 | 18 | Triangular |  |
| Days of hospitalization for ventilated patients | 19 | 15 | 32 | Triangular |  |
| Efficacy | Base | Low | High | Distribution | Source |
| Reduced days of hospitalization with remdesivir (from 15 days to 11 days) | 4 | 2 | 6 | Triangular | (7) |
| Mortality if those additional patients receive ICU care | 50% | 40% | 85% | Beta | (10) |
| Mortality if those additional patients do not receive ICU care | 90% | 85% | 100% | Beta | (10) |
| Mortality rate reduction for earlier stage (non-ventilated) patients due to remdesivir | 30% | 0% | 53% | Beta | (7) |
| Mortality rate reduction for later stage (ventilated) patients due to dexamethasone | 35% | 18% | 49% | Beta | (8) |
| Mortality rate reduction for earlier stage (non-ventilated) patients due to dexamethasone | 20% | 8% | 30% | Beta | (8) |
| Cost inputs | USD | Low | High | Distribution | Source |
| Health system operation cost in ICU per person day | \$1,128 | \$665 | \$1,172 | Gamma | (13) |
| Cost of remdesivir regimen (200 mg IV loading dose following by 100 mg IV daily for a maximum of 5 days) | \$330 | \$99 | \$330 | Gamma | (15) |
| Cost of dexamethasone regimen (6 mg PO / IV for up to 10 days) | \$31 | \$16 | \$47 | Gamma | (16) |

We performed one-way sensitivity analyses on all model parameters to describe the robustness of the primary results expressed as incremental cost per death averted to uncertainty in individual model parameters. Under our baseline assumptions, we also investigated whether and to what extent the benefit of a reduction in the length of ICU stay with remdesivir may differ by either assuming 1) capacity being breached for all months in all provinces vs 2) capacity never being breached. To explore the combined effect of uncertainty in our model parameters, we conducted a probabilistic sensitivity analysis (PSA). All model parameter values were randomly sampled over pre-specified distributions (Table 2) and 1,000 simulations were performed to produce cost-effectiveness acceptability curves. The model was constructed in Microsoft Excel using an Excel Visual Basic Macro to run the simulations.

## Results

In South Africa, there were a reported 3,318 public (1,178) and private (2,140) ICU beds available in total (11) and it was reported that only 70% (2,309 beds) were available for COVID patients as of 31 May 2020 (18). The estimated total health systems operation cost of COVID-19 related ICU care in South African excluding drugs during August 2020 and January 2021 is $83.9 million without remdesivir (standard care) and $71.4 million with remdesivir due to the reduction in required total ICU person days. In terms of drug costs, we estimated $983,000 required for remdesivir administered to non-ventilated patients, $67,000 for dexamethasone administered to ventilated patients, or $159,000 when dexamethasone is administered to all patients in the ICU across all six months.

Overall, the scenario with a combination of remdesivir and dexamethasone can save $11.5 million relative to standard care and avert a total of 1,111 deaths (Table 3). Of this, 473 deaths are averted due to remdesivir (assuming 30% mortality reduction due to remdesivir use), and 638 deaths averted due to dexamethasone on mortality. Similarly, the scenario with remdesivir to non-ventilated patients only can save $11.5 million relative to standard care but will result in fewer total deaths averted (473) compared to the scenario where both dexamethasone and remdesivir are used. The scenario with dexamethasone alone to ventilated and non-ventilated patients requires additional $159,048 but avert 1,146 deaths ($139 per death averted) relative to standard care. Therefore, should remdesivir become available, the use of remdesivir in combination with dexamethasone is recommended given the substantially greater cost saving for a similar health benefit.

**Table 3.** Total costs, health outcome, and incremental cost effectiveness ratios of COVID-19 treatment scenarios in South Africa from August 2020 - January 2021 (with 95% uncertainty ranges reported as the 2.5 ^th^ and 97.5^th^ percentiles of the corresponding distributions)

|  |  | Deaths averted |  |  | Cost per death averted |
| --- | --- | --- | --- | --- | --- |
| Scenarios | Total costs | Remdesivir | Dexamethasone | Total | *30% reduction in mortality of remdesivir* |
| Standard care | \$83,937,000<br>(\$43,612,000<br>\$104,632,000) | ref | ref | ref | ref |
| Dexamethasone to non-ventilated and ventilated patients | \$84,096,000<br>(\$44,260,000<br>\$104,497,000) | n/a | 1,146<br>(572 – 1,863) | 1,146<br>(572 – 1,863) | \$139<br>(-\$51, \$350) |
| Dexamethasone to ventilated patients only | \$84,003,000<br>(\$44,24,000<br>\$104,326,000) | n/a | 638<br>(227 – 1,140) | 638<br>(227 – 1,140) | \$104<br>(-\$36, \$336) |
| Dexamethasone to ventilated patients & Remdesivir to non-ventilated patients | \$72,465,000<br>(\$44,100,000<br>\$106,437,000) | 473*<br>(29– 1,293) | 638<br>(227 – 1,140) | 1,111<br>(239 – 2,027) | Cost saving |
| Remdesivir to non-ventilated patients only | \$72,399,000<br>(\$44,074,000<br>\$106,310,000) | 473*<br>(29– 1,293) | n/a | 473<br>(2 – 1,293) | Cost saving |

\*The number represents death averted by remdesivir assuming 30% reduction in mortality. It will be 9 (by decreasing the average length of stay of patients when ICU capacity is expected to be breached), if 0% reduction in mortality of remdesivir is assumed.

Our sensitivity analyses reveal that the uncertainty in the number of months ICU capacity is breached, and the efficacy of remdesivir and dexamethasone have the greatest impact on the ICER (Figure 1). For example, if ICU capacity is expected to be breached in all months and in all provinces, a scenario with remdesivir and dexamethasone combination will result in an additional drug costs of $9.3 million but will avert 6,562 deaths, resulting $1,421 per death averted; but if the ICU capacity is not expected to be breached at all, then the remdesivir and dexamethasone combination scenario can save up to $12 million (mainly due to the reduced total ICU days and health systems costs by remdesivir) and avert 659 deaths relative to standard care. If remdesivir reduces mortality by 30%, the scenario of remdesivir and dexamethasone combination will avert between 1,112 and 14,655 deaths ($636 per death averted) by the zero to the full (6) months across all provinces of ICU capacity being breached, respectively. The results of the PSA, displayed in the cost-effectiveness acceptability curves shows the probability of the intervention being cost-effective under different willingness to pay thresholds (Figure 2). For example, at a willingness to pay threshold of $1,000 per death averted, it is highly likely that the use of remdesivir and dexamethasone combined for critically ill COVID-19 patients in ICU will be cost effective (almost 100% of simulation estimates fell below the threshold) in various treatment scenarios compared to standard care in South Africa. The scenarios that include remdesivir show almost 50% of simulations fall below the cost saving threshold (i.e. incremental cost is less than 0) with more variability, mainly due to the uncertainty associated with the reduction in ICU days during non-breached months. Additionally, in the event that the mortality impact of remdesivir is 0% instead of 30%, the cost-effectiveness estimates do not change due to the substantial cost savings from reducing inpatient days.

**Figure 1.**
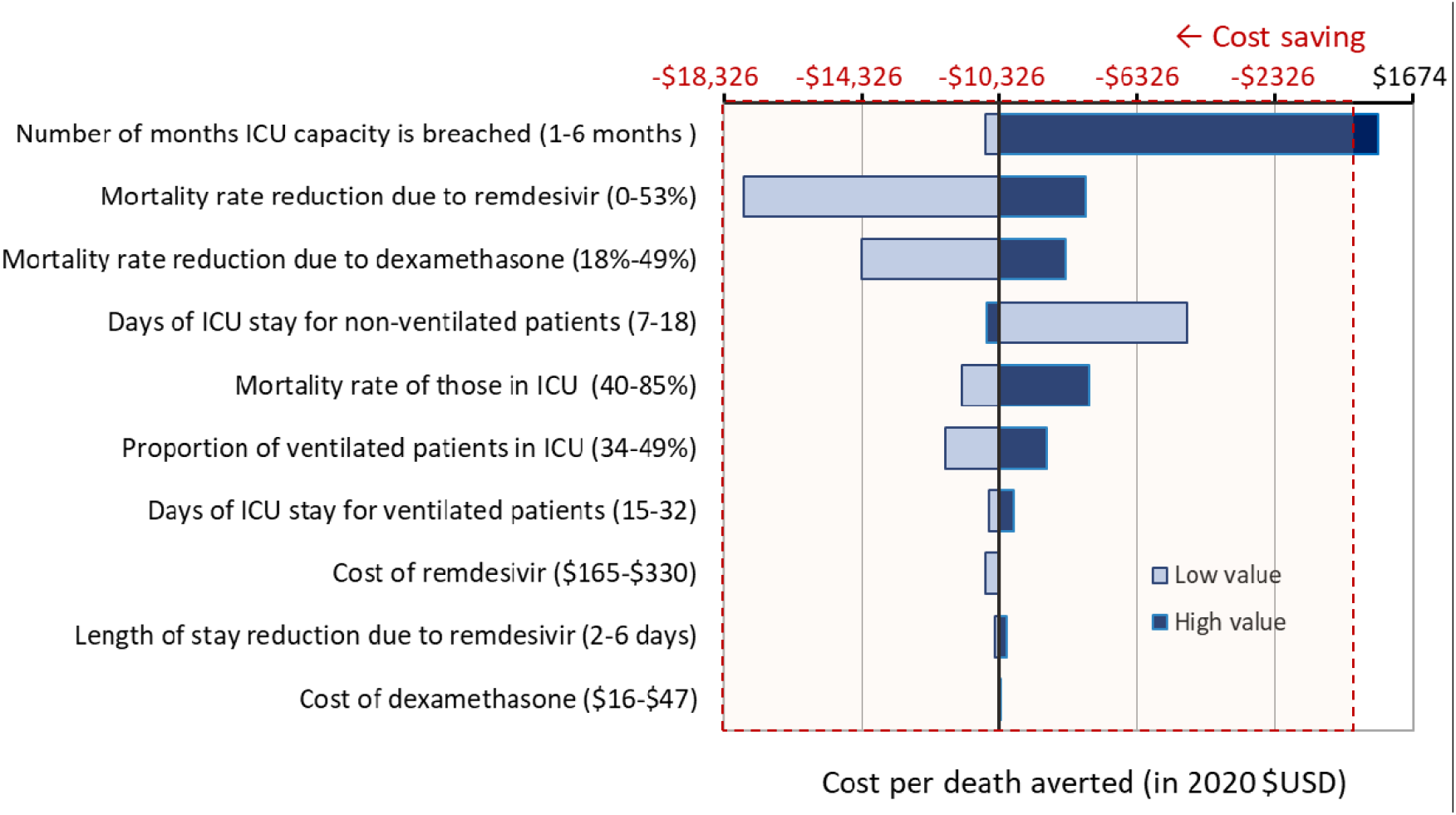
One-way sensitivity analyses of incremental cost effectiveness ratio comparison between a using remdesivir for non-ventilated ICU patients plus dexamethasone for ventilated patients compared to standard care (assuming 30% efficacy of remdesivir)

**Figure 2.**
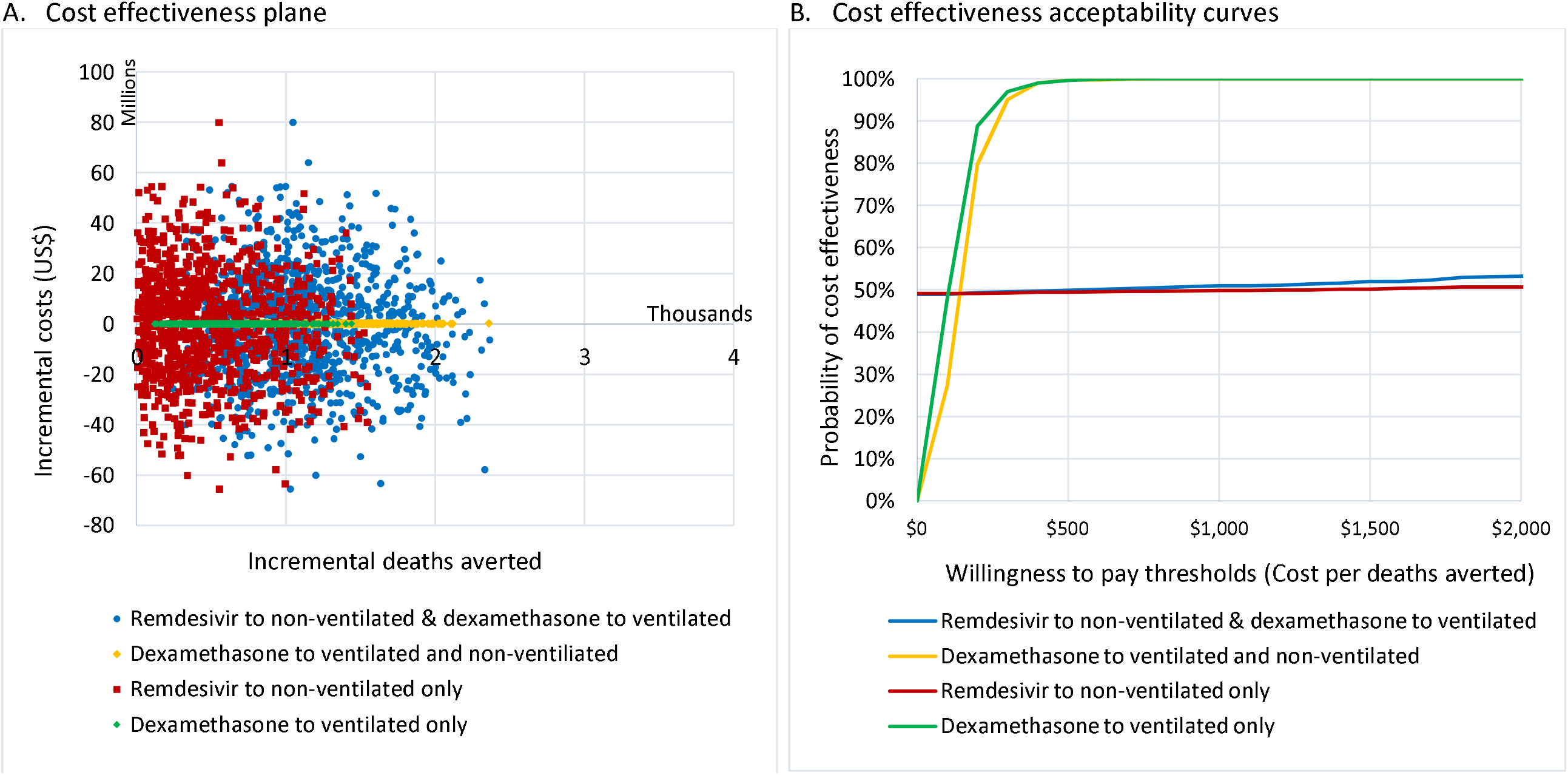
Cost-effectiveness plane and acceptability curves of the four dexamethasone and remdesivir treatment scenarios compared to standard care. *Variation of incremental cost in simulation output for the scenarios with dexamethasone is much narrower than the outputs for the scenarios including remdesivir because the incremental costs (relative to standard care) is only caused by drug costs (without change in length of ICU stay by the given number of ICU beds) and days by whereas the incremental cost of the scenarios with remdesivir (relative to standard care) is caused by both health system costs (due to the reduced length of ICU stay) and drug costs.

## Discussion

This analysis explores whether and to what extent remdesivir and dexamethasone might be considered cost-effective for critically ill COVID-19 patients in the context of a declining COVID-19 pandemic in South Africa. We find that a combination of remdesivir for non-ventilated patients and dexamethasone for ventilated patients is expected to maximize the number of deaths averted, and save the government of South Africa $11.5 million between August 2020 and January 2021. The results extend earlier findings regarding the mortality impact of remdesivir alone by reducing length of ICU stay and treating more COVID-19 patients in the ICU.(10) However, the months assumed for that early analysis included during the first peak of COVID-19 cases when resources were severely constrained. The present analysis is focused on the subsequent six months over which we expect the pandemic to continue to wane. This analysis demonstrates that remdesivir is likely to generate cost-savings and can offset the additional drug costs of dexamethasone to ventilated patients. This analysis therefore supports efforts to ensure access to remdesivir and dexamethasone even as COVID-19 epidemics wane – in order to save both lives and costs.

The number of deaths averted due to increasing ICU capacity in this analysis differ from our previous findings.(10) This is due to two factors. First, in this analysis we are assuming that only some patients in ICU receive remdesivir, instead of all patients. Second, the most recent set of model predictions from the NCEM project a waning epidemic in which ICU capacity is seldom breached. This differs previous projections that included the months of peak COVID-19 epidemic in South Africa (June-August 2020). This has resulted in much fewer deaths averted projected due to increasing ICU capacity.

Our analysis has several limitations. Firstly, the number of months ICU capacity is breached is influenced by the interplay between epidemic conditions and policy choices (which can affect ICU admission of other diseases).(19) As this will have different implications for costs and health outcomes in each scenario, it will be important to consider regional heterogeneity in epidemic conditions, health system capacity and willingness to pay in order to guide optimal treatment strategies. Second, we did not consider potential changes in the clinical course with disease progression nor change of distribution of disease severity among the patient population over time. Importantly, efficacy of remdesivir and dexamethasone in preventing mortality can be influenced by several factors – time of treatment initiation after symptom onset (20, 21), age (22), comorbidities(23), potential adverse events(24), and use of other medications(25, 26). As additional treatment options become available(27), it would also be important to collect more data on the duration of illness and its relationship to the outcome (both in terms of efficacy and safety)(24) and conduct more detailed analyses considering patient population characteristics, change of epidemic curves and local health system capacity, that can guide an optimal treatment strategy in a resource constrained setting. Third, our cost data does not include additional costs associated with adverse events management and also may not fully incorporate potential economies of scale due to increased volume of patients within a given ICU capacity. We also did not differentiate costs or financial implication between public and private hospitals. While this may change the point estimate of our results, it is unlikely to alter our conclusions. Continued efforts to develop clinical practice guideline (i.e. who to treat, where, how and when)(28) to reduce length of ICU stay, for patients in ICU with COVID-19 (29, 30) could further improve clinical benefits and cost-effectiveness. Finally, given that cost-savings observed with remdesivir is driven by months when ICU capacity is not breached, the value for money of remdesivir may increase during the next phase of the pandemic as new cases decline and the pressure on ICUs eases. Given great uncertainty in making long-term projections into 2021, the time horizon for this study was limited to latest projections from the NCEM.

## Conclusion

To conclude, treatment with remdesivir for non-ventilated patients and dexamethasone for ventilated patients will maximize lives saved and is predicted to save $11.5 million between August 2020 and January 2021. The results were strongly driven by the efficacy of the drugs, and the reduction of ICU-time required for patients treated with remdesivir. We therefore recommend that future studies collect additional data on mortality impact stratified by disease severity and care condition in South Africa to further refine cost-effectiveness analyses and guide implementation. This analysis strongly supports ensuring access to remdesivir and dexamethasone to save costs and lives in South Africa.

## Data Availability

Data consisted of model output and parameters from existing literature. Data available upon request from corresponding author.

